# Characterising heterogeneity and sero-reversion in antibody responses to mild SARS⍰CoV-2 infection: a cohort study using time series analysis and mechanistic modelling

**DOI:** 10.1101/2020.11.04.20225920

**Authors:** C Manisty, TA Treibel, M Jensen, A Semper, G Joy, RK Gupta, T Cutino-Moguel, M Andiapen, J Jones, S Taylor, A Otter, C Pade, JM Gibbons, WYJ Lee, M Jones, D Williams, J Lambourne, M Fontana, DM Altmann, RJ Boyton, MK Maini, A McKnight, T Brooks, B Chain, M Noursadeghi, JC Moon, on behalf of COVIDsortium Investigators

**Affiliations:** Institute of Cardiovascular Sciences, University College London, London, UK; Barts Heart Centre, St Bartholomew’s Hospital, Barts Health NHS Trust, London, UK, London, UK; National Infection Service, Public Health England, Porton Down, UK; Division of Infection and Immunity, University College London, London, UK; Department of Virology, Barts Health NHS Trust, London, UK; Centre for Cardiovascular Medicine and Devices, William Harvey Research Institute, Queen Mary University of London, London, UK; Blizard Institute, Barts and the London School of Medicine and Dentistry, Queen Mary University of London, London, UK; Wolfson Institute of Preventive Medicine, Barts and the London School of Medicine and 24 Dentistry, Queen Mary University of London, London, UK; MRC Unit for Lifelong Health and Ageing, University College London, London, UK; Department of Medical Epidemiology and Biostatistics, Karolinska Institutet, Stockholm, Sweden; Department of Infection, Barts Health NHS Trust, London, UK; Royal Free London NHS Foundation Trust, London, UK; Division of Medicine, University College London, London, UK; Department of Immunology and Inflammation, Imperial College London, London, UK; Department of Infectious Disease, Imperial College London, London, UK

**Author notes:** **Correspondence** Professor Mahdad Noursadeghi, Division of Infection and Immunity, University College London. These authors made an equal contribution (M Noursadeghi, JC Moon).

## Abstract

**Background:** SARS-CoV-2 serology is used to identify prior infection at individual and at population level. Extended longitudinal studies with multi-timepoint sampling to evaluate dynamic changes in antibody levels are required to identify the time horizon in which these applications of serology are valid, and to explore the longevity of protective humoral immunity.

**Methods:** Health-care workers were recruited to a prospective cohort study from the first SARS-CoV-2 epidemic peak in London, undergoing weekly symptom screen, viral PCR and blood sampling over 16-21 weeks. Serological analysis (n=12,990) was performed using semi-quantitative Euroimmun IgG to viral spike S1 domain and Roche total antibody to viral nucleocapsid protein (NP) assays. Comparisons were made to previously reported pseudovirus neutralising antibody measurements.

**Findings:** A total of 157/729 (21.5%) participants developed positive SARS-CoV-2 serology by one or other assay, of whom 31.0% were asymptomatic and there were no deaths. Peak Euroimmun anti-S1 and Roche anti-NP measurements correlated (r=0.57, p<0.0001) but only anti-S1 measurements correlated with near-contemporary pseudovirus neutralising antibody titres (measured at 16-18 weeks, r=0.57, p<0.0001). By 21 weeks’ follow-up, 31/143 (21.7%) anti-S1 and 6/150 (4.0%) anti-NP measurements reverted to negative. Mathematical modelling suggested faster clearance of anti-S1 compared to anti-NP (median half-life of 2.5 weeks versus 4.0 weeks), earlier transition to lower levels of antibody production (median of 8 versus 13 weeks), and greater reductions in relative antibody production rate after the transition (median of 35% versus 50%).

**Interpretation:** Mild SARS-CoV-2 infection is associated with heterogenous serological responses in Euroimmun anti-S1 and Roche anti-NP assays. Anti-S1 responses showed faster rates of clearance, more rapid transition from high to low level production rate and greater reduction in production rate after this transition. The application of individual assays for diagnostic and epidemiological serology requires validation in time series analysis.

**Funding:** Charitable donations via Barts Charity

**Research in context:** *Evidence before this study:* We searched PubMed, medRxiv, and bioRxiv for [“antibody” OR “serology”] AND [“SARS-CoV-2” OR “COVID-19”]. The available literature highlights widespread use of serology to detect recent SARS-CoV-2 infection in individual patients and in population epidemiological surveys. Antibody to virus spike protein S1 domain is widely reported to correlate with neutralising antibody titres. The existing assays have good sensitivity to detect seroconversion within 14 days of incident infection, but the available longitudinal studies have reported variable rates of decline in antibody levels and reversion to undetectable levels in some people over 3 months. High frequency multi-time point serology data for different antibody targets or assays in longitudinal cohorts from the time of incident infection to greater than 3 months follow up are lacking.

*Added value of this study:* We combine detailed longitudinal serology using the Euroimmun anti-S1 and Roche anti-nucleocapsid protein (NP) assays in 731 health care workers from the time of the first SARS-CoV-2 epidemic peak in London, UK. In 157 seroconverters (using either assay) we show substantial heterogeneity in semiquantitative antibody measurements over time between individuals and between assays. Mathematical modelling of individual participant antibody production and clearance rates in individuals with at least 8 data points over 21 weeks showed anti-S1 antibodies to have a faster clearance rate, earlier transition from the initial antibody production rate to lower rates, and greater reduction in antibody production rate after this transition, compared to anti-NP antibodies as measured by these assays. As a result, Euroimmun anti-S1 measurements peaked earlier and then reduced more rapidly than Roche anti-NP measurements. In this study, these differences led to 21% anti-S1 sero-reversion, compared to 4% anti-NP sero-reversion over 4-5 months.

*Implications of all of the available evidence:* The rapid decline in anti-S1 antibodies measured by the Euroimmun assay following infection limits its application for diagnostic and epidemiological screening. If generalisable, these data are consistent with the hypothesis that anti-S1 mediated humoral immunity may not be sustained in some people beyond the initial post-infective period. Further work is required to understand the mechanisms behind the heterogeneity in antibody kinetics between individuals to SARS-CoV-2. Our data point to differential mechanisms regulating humoral immunity against these two viral targets.

## Introduction

Detection of antibodies to SARS-CoV-2 is key to establishing the prevalence of infection in the population, and hence tracking the progress of the pandemic, and may be used to diagnose past infection in individual patients. Moreover, antibody to envelope spike protein may contribute to protective immunity.^1-4^ Interpretation of cross-sectional serology is critically dependent on understanding the dynamics of the antibody response, and how this might vary for different viral target antigens, in different assays and between individuals.

Numerous studies have shown that individuals with a confirmed polymerase chain reaction (PCR) diagnosis of SARS-CoV-2 develop IgM, IgA and IgG against the spike protein S1 domain and nucleocapsid protein (NP) within 2 weeks of symptom onset, which remain detectable following initial viral clearance.^5-7^ Early data suggest antibody levels correlate with disease severity.^8,9^ Antibody responses to other human coronaviruses decay over time with regular reinfection events, which has caused concern that SARS-CoV-2 immunity following natural infection may be short-lived, leading to risk of re-infection and making the possibility for achieving herd immunity through natural infection unrealistic.^10-12^ The data for SARS-COV-2 remains conflicting, with some longitudinal serological studies suggesting rapid antibody decline, while others have shown much greater persistence.^3,13-22^ These vary by cohort (hospital, symptomatic only, PCR positive, community), assay (quality-assured, antigen target), sampling granularity and follow-up period.

We present a detailed temporal analysis of circulating antibody using two widely used semi-quantitative commercial assays to detect either anti-S1 or anti-NP in a cohort of hospital health care workers in a prospective longitudinal multi-centre cohort study with high frequency serial sampling over 16-21 weeks during the first epidemic wave in London, UK. We assessed concordance between assays and the determinants of inter-individual heterogeneity in antibody responses by testing associations with clinical and demographic variables. Finally, we applied mathematical modelling to infer the fundamental mechanisms that may underpin changes to antibody levels over time.

## Methods

### Study design and participants

The study was approved by a UK Research Ethics Committee (South Central - Oxford A Research Ethics Committee, reference 20/SC/0149). The details of participant screening, study design, sample collection, and sample processing are previously published and registered on ClinicalTrials.gov, NCT04318314.^23^ Briefly, hospital healthcare workers (HCWs) self-declared fit to attend work were recruited to an observational cohort study consisting of questionnaires and biological sample collection at baseline and over 16 weekly-follow-up visits. Those who were unable to attend follow-up visits were consulted remotely to enable capture of information regarding possible exposures and symptoms. The baseline questionnaire included demographic data, medical history and exposures, alongside detailed information regarding the nature and timing of self-reported symptoms over the preceding 3 months.^23^ Follow-up weekly questionnaires included data on new symptoms and changes to occupational and community risk factors, or results of tests conducted outside of the study. Symptoms were classified as follows: ‘case-defining’ (fever, new dry cough or a new loss of taste or smell; which have been shown to predict COVID-19 positivity with high specificity), ‘non-specific (symptoms other than case-defining symptoms), or asymptomatic (no symptoms reported throughout the study period or in the three preceding months).

An initial cohort of 400 HCWs was recruited from St Bartholomew’s Hospital, London, UK in the week of lockdown in the United Kingdom (between 23^rd^ and 31^st^ March 2020); Cohort 1. Recruitment was subsequently extended to include additional participants from multiple sites between 27^th^ April 2020 and 7^th^ May 2020 (Cohort 2). This included St Bartholomew’s Hospital (n=101), NHS Nightingale Hospital (n=10), and the Royal Free NHS Hospital Trust (n=220). Data collection therefore extended over 21 weeks from baseline recruitment of Cohort 1 (the day of UK lockdown) to completion of 16-week follow-up of Cohort 2. Towards the end of the study period for Cohort 2 (follow-up weeks 12-15) following the decline in community infection rates, the frequency of blood sampling was reduced to twice per month rather than weekly, in order to improve tolerability to participants.

### Procedures

Nasal RNA stabilizing swabs for molecular testing for SARS-CoV-2 were acquired at baseline and weekly. RT-PCR was performed on nasal swabs using Roche cobas® SARS-CoV-2 test. SARS-CoV-2 antibody testing was performed in a single laboratory at Public Health England on all available serum samples from baseline and follow-up visits using two commercial assays according to manufacturers’ protocols. These were the Euroimmun anti-SARS-CoV-2 enzyme-linked immunosorbent assay (ELISA) (IgG) targeting IgG specific for the SARS-CoV-2 S1 antigen, and the Roche Elecsys Anti-SARS-CoV-2 electrochemiluminescence immunoassay (ECLIA) that detects antibodies (including IgG) directed against the SARS-CoV-2 nucleocapsid protein (NP).^24-27^

The Euroimmun ELISA was performed using a Stratec Gemini automated microplate processor as previously described.^28^ Raw optical density (OD) readings were adjusted by calculating the ratio of the OD of the control or participant sample divided by the OD of the assay calibrator. A ratio ≥1.1 was used as the threshold for a positive result as per manufacturer’s instructions.^24^ A ratio of 11 was used as the upper threshold of the dynamic range, as the assay saturated above this point. The Roche ECLIA was performed using the Roche cobas^®^ e801 immunoassay analyser analyzer.^26^ Results are expressed as a cut-off index (COI), calculated by the analyser software as the electrochemiluminescence signal obtained from the patient sample divided by the lot-specific cut-off value.^25^ A COI≥1 was used as the threshold for a positive result as per manufacturer’s instructions. Across their dynamic range, the semi-quantitative indices of both assays approximate to a linear relationship with antibody levels (Supplementary Figure 1). We have previously reported quantitation of pseudovirus neutralising antibody (nAb) titres in 70 seropositive HCW from this cohort at 16-18 weeks of follow up.^29^

### Statistical analysis

In descriptive analyses, Spearman’s rank correlation coefficients were calculated for all paired assay values across the study period. Univariable associations of demographics, symptoms and exposures with serostatus (seropositivity by one or both antibody assays at any timepoint) were assessed using logistic regression. Univariable and multivariable associations of characteristics (including age, sex, ethnicity and case-defining symptom status) with peak antibody levels were also quantified using linear regression for anti-N IgG/IgM and anti-S1 IgG.

We also performed univariable and multivariable survival analyses to assess whether participant characteristics (including age, sex, ethnicity, case-defining symptom status and peak anti-S1) were associated with time to sero-reversion for the anti-S1 assay. For this analysis, we included all participants who seroconverted on the anti-S1 assay at any point during follow-up. We assumed synchronous onset of infection for these individuals by indexing the start time for the survival analysis as the first week of Cohort 1 study enrolment (the week of UK lockdown). Anti-S1 sero-reversion was defined as a negative test in the last assay performed during follow-up for each participant. Participants who sero-reverted exited the survival analysis on the first week where a negative test (following an earlier positive test) was recorded; those who did not sero-revert were censored on the week of their last available anti-S1 serology result. Analyses were conducted in R (version 3.6.3) and Stata Statistical Software version 16 (College Station, TX, USA).

### Mechanistic mathematical modelling of antibody production

Circulating antibody levels are determined by the balance between rates of production and clearance. We represented antibody production by a simplified discrete mechanistic model captured by equation 1, with time indexed to calendar weeks from initiation of UK lockdown (as described for the survival analyses). We incorporated a production rate (AbPr) and an antibody turnover (clearance rate, r). Antibody production was simplified to two phases, an initial high rate (AbPr1) followed by a switch to a lower rate (AbPr2), after a time t_stop. Since the assay units of antibody concentration are arbitrary and are not comparable between assays, the value of AbPr 1 is also arbitrary, and serves only to scale the model output to the scale of the data. AbPr2 is expressed as a proportion of AbPr1. The rate of clearance r can be directly calculated from the half-life, which was allowed to vary between 1 week and 4 weeks (the latter equivalent to the known turnover rate of free IgG). An important emerging feature of the model is that the time to plateau (peak) is determined only by the clearance rate (Supplementary Figure 2), and not by the rate of production AbPr1. Furthermore, any subsequent fall from the peak must reflect a corresponding decrease in AbPr. Hence the model assumption that AbPr2<-AbPr1.

Equation 1: Ab_t_ = Ab_t-1_ + AbPr – Ab_t-1_ * (1 – e^-rt^), where t is time in weeks, AbPr = AbPr_1_ for 1 < t < t_stop or AbPr_2_ for t_stop < t < t_end; and AbPr2<AbPr1, and r = log(2)/half_life.

The levels of anti-S1 or anti-NP antibody in blood were compared to the model, over a range of the parameters (AbPr1, AbPr2 as a proportion of AbPr1, r and t_stop) by calculating the root mean square distance between data and model output, and the parameter set with the minimum distance was selected. In our primary analysis, we restricted mathematical modelling to seropositive participants with ≥8 antibody data points (N=92 for anti-S1, N=86 for anti-NP, Supplementary Figure 3). In sensitivity analysis, we further restricted the modelling to seroconverters in whom the first (baseline) sample was negative.

## Results

### Study population

The study population has previously been published^23^ and are summarised in Supplementary Table 1. Briefly, this comprised 731 HCWs (median age 35 IQR 28 years, 33.0% male). Fourteen withdrew (two with no samples obtained). Average weekly attendance was 61% with median 10 (IQR 6) visits per participant. 20.2% were doctors, 31.2% were nurses, 27.5% were allied healthcare professionals, 21.1% were others including administrative and clerical. 22.6% worked in an intensive care unit or emergency department setting. Co-morbidities were relatively low (18.1% smokers, 12.6% BMI>30kg/m^2^, 10.7% with asthma, 7.3% with hypertension, 2.1% with diabetes mellitus, 1.2% with rheumatological disease, 0.8% with cancer). 62.5% of participants were white, 37.5% non-white with 5.6% of black ethnicity), and 47.6% reported a mean household size of ≥3 people. Exposure at baseline to contacts with confirmed COVID-19 was high (42.7% to patients, 29.5% to colleagues, and 1.1% to household members with confirmed COVID-19) and 25.6% were exposed to aerosol generating procedures.

### Seroconversion to SARS-CoV-2

In cohort 1 (recruited from 23^rd^ March; day of UK lockdown), 28/396 (7.1%) had a positive nasal PCR at baseline, with rates falling rapidly in the subsequent four weeks, Figure 1.^30^ In cohort 2 (recruited from 27^th^ April, 5 weeks after UK lockdown and study start) 3/331 (0.9%) had positive nasal PCR swabs at their first study visit. There were no positive PCR swabs across either cohort by 6 weeks after UK lockdown (after 1st May 2020). The cumulative PCR positivity rate was 6.6% (48/729).

**Figure 1.**
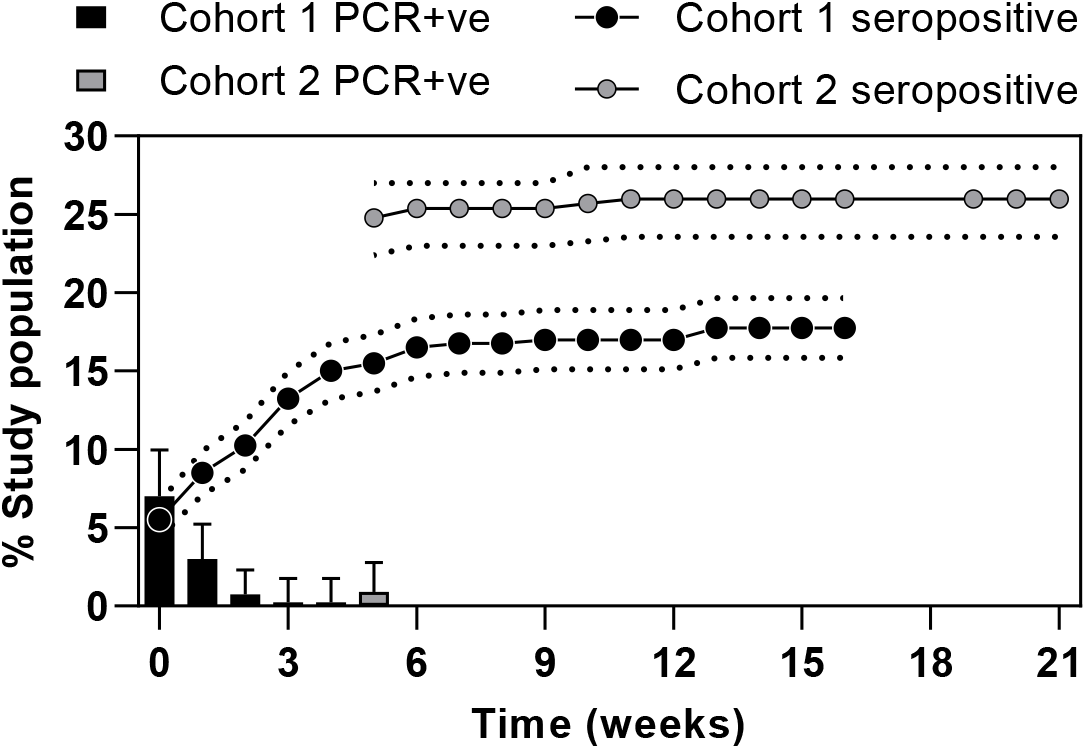
Longitudinal infection with SARS-COV-2 over 21 weeks across 731 healthcare workers. Results from testing for SARS-CoV-2 by cohort showing weekly PCR percentage positivity (weekly results, 95% CI) and seropositivity (cumulative percentage using combined anti-S1 IgG and anti-NP IgM/IgG, standard error).

Serology testing used both assays on all subjects at all timepoints giving a total of 12990 tests and median of 10 paired assays per individual (Supplementary Figure 3). Baseline seropositivity (by either assay) in cohort 1 was 22/399 (5.5%), rising to 17.8% by study completion and 82/330 (25%) rising to 26.1% for cohort 2 (which started 5 weeks later). Overall, 157 of 729 (21.5%) had at least one seropositive result. Consistent with the lack of nasal swab PCR detection of incident infection by 6 weeks after UK lockdown, 98% of cumulative seroconversions were evident by 7 weeks after lockdown. Of the 48 participants in whom incident infection was detected by a positive nasal swab PCR at any time point, 44 had subsequent blood tests of which 42/44 (95.4%) became seropositive in at least one assay. Subsequent anti-S1 seropositivity at any timepoint was lower following a positive PCR result than anti-NP seropositivity (86.4% versus 93.2% respectively), although the time interval to seroconversion was faster (median 2.5 versus 3.0 weeks). Across all samples, binary outcomes at the manufacturers’ predefined thresholds for each assay were concordant in 96.9% (6276/6476 samples), but this reduced to 82.7% (953/1153) concordance in those samples where at least one of the two assays were positive.

Of participants who were seropositive at any timepoint, 43.9% reported case-definition symptoms during the study, 24.8% non-case definition symptoms and 31.2% were completely asymptomatic. Only two study participants were hospitalised, neither required ventilatory support or died. In univariable analysis, there was no association between age or sex and seropositivity, but risk was higher in participants of Black ethnicity (odds ratio 2.61 [1.36, 4.98], p=0.004, Supplementary Figure 4). Risk of infection could not be explained by baseline co-morbidities or clinical roles, although HCWs based in ICU had lower rates than others (OR 0.52 [0.30, 0.90], p=0.02). Reported exposure to household members (baseline plus follow-up) with COVID-19 was the strongest association with infection (OR 11.36 [2.27, 56.87], p=0.003). In contrast, reported exposure to SARS-CoV-2 positive patients or colleagues did not influence infection rates.

### Longitudinal serology to SARS-CoV-2

Peak antibody measurements were highly variable between seropositive individuals across the cohort (coefficient of variation 77.89% for anti-NP and 54.09% for anti-S1, Figure 2). Despite infections being mild, 7.7% participants had values at the threshold upper limit in the Euroimmun anti-S1 assay. We extended our previous exploration of associations with peak antibody measurements in subset of the cohort^29^ to the full cohort of seropositive participants (Supplementary Table 1-3). In multivariable analyses, there was a modest positive association of increasing age with peak anti-S1 antibody measurements (beta coefficient per year increase 0.05; 95% CI 0.01 to 0.09; p=0.021), but not with anti-NP (beta coefficient 0.50; 95% CI −0.22 to 1.2; p= 0.2). Black, Asian and minority ethnic groups were associated with higher peak anti-NP responses (beta coefficient 22; 95% CI 4.9 to 39; p= 0.012), with a weaker association for peak anti-S1 (beta coefficient 1.0; 95% CI –0.04 to 2.0; p= 0.058). We found no association of peak antibody measurements with sex, or case-defining symptom status.

**Figure 2.**
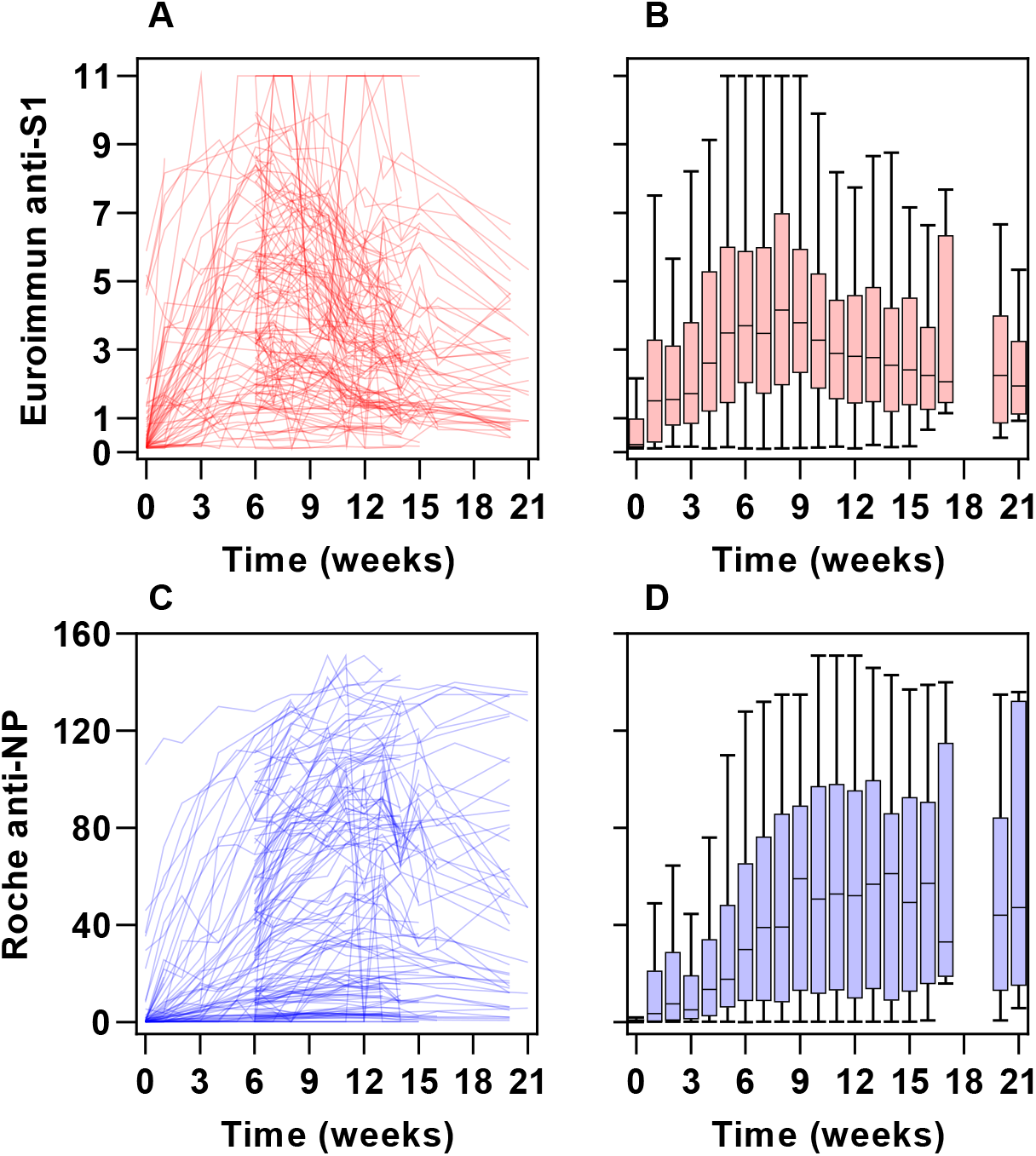
Longitudinal antibody responses across all timepoints in participants seropositive at any timepoint. Individual participant data (left) and time ordered aggregate data (right) for Euroimmun anti-S1 antibody assay (**A-B**) and Roche combined anti-NP antibody assay (**C-D**) showing the heterogeneity in antibody responses between individuals and the differences in antibody kinetics between assays, with an earlier peak and decline in anti-S1 compared with anti-NP antibodies.

Peak antibody measurements using the two assays correlated (r=0.57, p<0.001) (Supplementary Figure 5). The correlation between ranked antibody indices stratified by time interval from the start of the study, revealed a shift from more highly ranked anti-S1 antibodies in the first 6 weeks to more highly ranked anti-NP antibodies in the last 8 weeks (Supplementary Figure 6A), and the ratio of anti-S1:anti-NP antibody measurements trended downwards over time (Supplementary Figure 6B). This analysis was consistent with different temporal profiles in circulating antibody levels to these two targets. Accordingly, among seropositive participants, time ordered aggregate data for each assay showed anti-S1 antibody indices to reach a peak and then fall more rapidly than the anti-NP antibody indices (Figure 2). By the end of the study 31/143 (21.7%) with positive Euroimmun anti-S1 serology had reverted to negative, in comparison to 6/150 (4%) of those with positive Roche anti-NP serology.

We compared previously reported neutralising antibody (nAb) titres in all seroconverters who attended for an additional blood sample (N=70) at 16-18 weeks of follow up^29^ to the highest anti-S1 and anti-NP assay measurements in the present study in 54/70 participants for whom we had data from within two weeks of nAb measurements. The inhibitory concentration (IC)50 titres showed significant correlation to ranked anti-S1 antibody indices (r=0.57, p<0.0001) but not to anti-NP antibody indices at near-contemporary time points (Figure 3).

**Figure 3.**
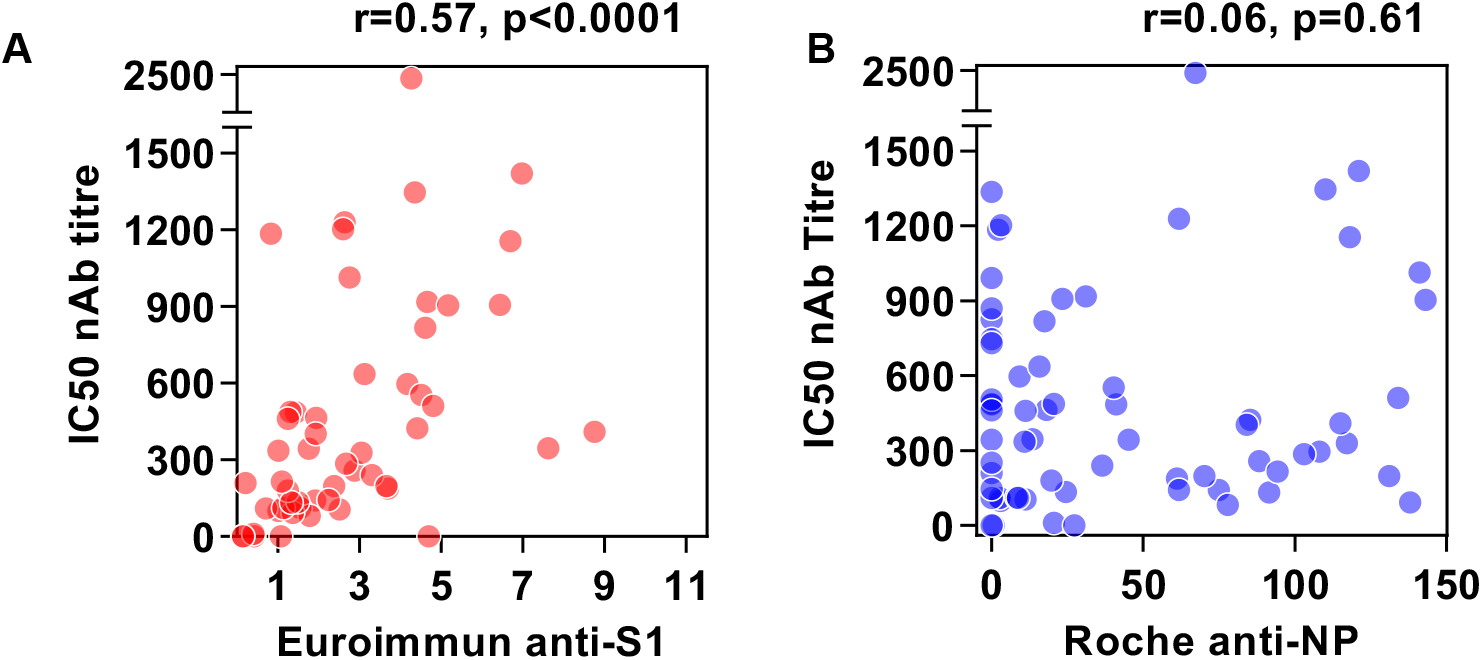
Correlation of anti-S1 and anti-NP antibody measurements with neutralising antibody titres at 16-18 weeks. Comparison of neutralising antibody (nAb) titres represented as 50% inhibitory concentrations (IC) with Euroimmun anti-S1 levels **(A)** and Roche anti-NP levels **(B)** in 54 participants with nAb measurements and near-contemporaneous (±2 weeks) Euroimmun and Roche serology. R and p values by Spearman rank correlations.

In univariable and multivariable survival analyses among participants who had a positive anti-S1 assay, higher peak anti-S1 responses were associated with longer time to sero-reversion (hazard ratio 0.42 per unit increase when adjusted for age, sex, ethnicity and case-defining symptom status; 95% CI 0.30 to 0.58; p<0.001; Supplementary Table 4). Age, sex, ethnicity and presence/absence of case-defining symptoms were not associated with time to sero-reversion.

### Mathematical modelling of kinetics of circulating anti-S1 and anti-NP antibodies

We sought to obtain further insight into these underlying processes by fitting a mathematical model to the antibody data. We first fitted the model to the median of all the data for all individuals who were seropositive at any time point, assuming approximately synchronous infections coincident with the peak epidemic transmission at the start of the study. The best fit models for the anti-S1 and anti-NP data were clearly distinct. The inferred rate of clearance of S1 antibodies was faster than that of NP antibodies, and the switch to a lower antibody production rate occurred sooner and reduced by a greater extent (Figure 4A-B).

**Figure 4.**
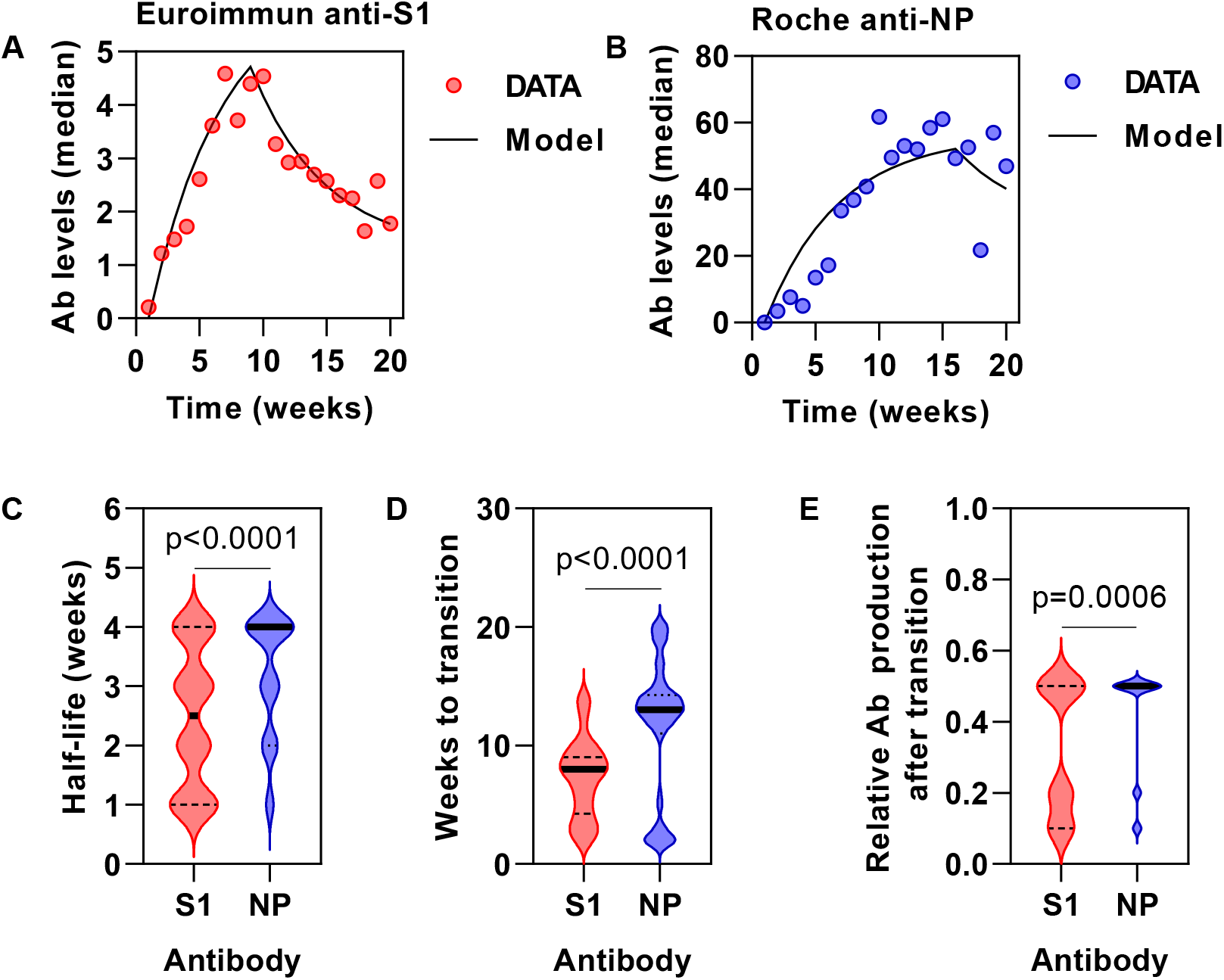
Mathematical modelling of kinetics of circulating anti-S1 and anti-NP antibodies. Model fit to aggregate (median) data from all seropositive participants for anti-S1 **(A)** and anti-NP **(B)** assays. Best fit model parameters for individual seropositive participants for half-life of antibody clearance **(C)**, time to transition point of lower antibody production **(D)** and relative reduction in antibody production following this transition point **(E)**. Horizontal lines **(C-E)** show median and interquartile range.

We noted that individual antibody response profiles were heterogenous in magnitude and dynamics (Figure 2A). We therefore repeated our analysis for each subject individually (Supplementary Figure 6), and derived the model parameters (Figure 4C-E). As anticipated, the best fit model parameters across the cohort were highly heterogenous. However, clear differences could be observed between the anti-S1 and anti-NP antibody responses, reflecting the same hierarchy as we observed when fitting the median antibody data. Thus, anti-S1 antibodies have a shorter half-life (median 2.5 weeks, 95% CI 2-3) than anti-NP antibodies (median 4 weeks, 95% CI 3-4). Production switches to a lower rate more quickly (median 8 weeks, 95% CI 7-8 versus 13 weeks, 95% CI 13-14), and to a relatively lower level (median 0.35, 95% CI 0.2-0.5 versus 0.5, 95% CI 0.05-0.5) with anti-S1 than with anti-NP antibodies, respectively. In general, there appeared to be greater inter-individual heterogeneity in the best fit model parameters for the Euroimmun anti-S1 antibody profiles than for the Roche anti-NP antibody profiles (Figure 4C-E). There were no strong associations between model parameters (half-life of antibody clearance, time to lower production rate and level of reduction) for either assay with age, sex, ethnicity or symptoms.

In order to exclude the potential confounding effects of capturing individuals only after antibody production was well established, we repeated the analysis using only those who were seronegative at the first available timepoint. The results were qualitatively the same for this smaller sub-group (Supplementary Figure 8).

## Discussion

We report a detailed time series analysis of circulating antibodies to SARS-CoV-2 S1 and NP proteins following asymptomatic or pauci-symptomatic infection, using two widely adopted commercial assays. Antibody measurements were highly heterogeneous between seropositive individuals, however both assays show initial high sensitivity for incident (even asymptomatic) infections following PCR detection of virus on nasal swabs, consistent with previous evaluations of these tests.^31 26,27^ Peak antibody index measurements in each assay were significantly correlated, suggesting both antigenic targets were similarly immunogenic. By 16-21 weeks after the peak of the epidemic wave in London, more than one in five individuals who had developed positive serology to the virus spike protein had sero-reverted by the Euroimmun anti-S1 assay. In contrast, reduction in Roche anti-NP measurements to subthreshold levels was evident in less than one in twenty. These findings have potentially important implications. First, it reveals that epidemiological seroprevalence surveys may be biased by antibody decay over this time horizon and may substantially underestimate incident infections. Second, given that anti-S1 antibodies correlate with protective neutralising antibodies, the extent of the reduction in circulating levels of anti-S1 suggests that antibody-mediated protective immunity in some individuals may be short-lived following asymptomatic or pauci-symptomatic infection.

Semi-quantitative antibody measurements over the initial five months following infection revealed differential kinetics for the two assays. Anti-S1 antibodies reached peak levels before anti-NP antibodies, but also showed a more rapid decline. It is interesting to speculate whether differential profiles of antibody assays can be exploited to estimate the time of infection. We used mathematical modelling to evaluate the determinants of the kinetic profiles and showed that the differential time to peak antibody level is dependent on differential clearance rates. Our model is consistent with a median half-life of 4 weeks for anti-NP antibodies consistent with long established estimates for circulating IgG. The median half-life of anti-S1 antibodies was significantly less, at 2.5 weeks. The subsequent fall in antibody levels reveals a transition in antibody production to a lower level. Our model estimated that this transition occurred at a median of 8 weeks for anti-S1 antibodies compared to a median of 14 weeks for anti-NP antibodies. Finally, we sought to derive the relative rate of antibody production after the transition. For both antibodies, this reduced to at least 50% of antibody production rates before the transition, but a substantial proportion of individuals exhibited significantly greater reduction of anti-S1 antibody production. The combination of lower antibody production rate and the natural clearance of antibody resulted in levels falling below the detection threshold of the assay in a significant proportion of the study cohort. In multivariable analysis, only peak antiIZIS1 measurements were associated with shorter time to anti-S1 sero-reversion.

The durability of antibodies to specific antigenic targets is highly variable after different viral infections and the factors which determine these are poorly understood. Several hypotheses merit investigation in future work. If the surface exposed domains of the SARS-CoV-2 spike protein have greater propensity to form immune complexes, increased rates of antibody clearance via immune complex formations^32^ may contribute to the shorter half-life of anti-S1 antibodies. The transition to lower levels of antibody production may represent the switch from antibody production from short lived plasmablasts to long lived plasma cells.^33^ Importantly, immune complexes are also known to regulate antibody production via inhibitory Fc receptors on plasma cells.^34^ Therefore, immune complex formation may also contribute to lower levels of anti-S1 antibody production after the transition. Alternatively, there may be differences in the relative contribution of short-lived extrafollicular memory B cells versus long-lived plasma cells to the antibody responses against these two antigens. We found a stronger T cell response at 16-18 weeks to whole NP than whole spike antigen in this cohort^29^; future studies should test whether NP-specific T follicular helper cells are better equipped to support an efficient germinal centre reaction resulting in more long-lived plasma cells for NP than spike protein. Alternatively, the better durability of anti-NP antibodies could relate to differences in maintenance of their cognate antigen, for example on follicular dendritic cells.

A key strength of our study is that the start at the time of first epidemic peak in London, UK, allowed the time of incident infection to be estimated accurately (69.0% of cohort 1 recruited prior to seroconversion) and the data were not confounded by prior exposures or vaccine trials. In addition, the focus on asymptomatic and pauci-symptomatic infection is representative of the vast majority of incident infections. Our finding that almost one in three were completely free from symptoms despite detailed weekly contemporaneous data collection to reduce recall bias is consistent with other estimates of rates of asymptomatic infection.^22^ Serology was assessed weekly with a median of 10 samples per participant over 16 weeks and, with almost 13,000 validated antibody assays performed, provide to our knowledge the most granular longitudinal data currently available at this scale.

Previous reports of the longevity of circulating antibodies to SARS-CoV-2 vary. Direct comparisons are undermined by the use of different assays targeting distinct antigens, differences in study population demographics, severity of illness, sampling frequency and duration of follow up. Two themes have emerged. First, that there is a detectable rate of reversion of seropositive individuals to becoming seronegative over 3 to 6 months, consistent with our findings.^3,18,19^ Some reports have sought to predict the time to sero-reversion with multilevel models to estimate the decay rate.^20,35^ Our analysis extends these analyses significantly by combining unprecedented high frequency sampling and mathematical modelling to provide dynamic estimates of production and clearance rates that determine the overall levels of circulating antibody. Second, there are frequent reports of an association between the clinical severity of infection with magnitude of initial antibody responses and the longevity of circulating antibody titres.^15,35^ Whether, this explains recent studies that show sustained levels in hospitalised patients over 3 to 6 months will require further evaluation.^2,36^

Our study has important limitations. Our time series analysis was limited to individual semiquantitative assays for each antigenic target. Direct comparison of antibody levels was not possible due to differences in dynamic range of these assays and their co-linearity. At present, results from any single assay are not generalisable. Moreover, these assays did not provide any differential assessment of antibody subclasses, which may exhibit differential kinetics. The significant correlation between near-contemporary Euroimmun anti-S1 measurements and functional pseudovirus nAb titres increased confidence in our assessment of anti-S1 levels, and is in line with data using live virus micro-neutralisation.^37^ Nonetheless, the correlation coefficient was modest suggesting that Euroimmun anti-S1 measurements do not explain all humoral neutralising activity. Moreover, emerging data on T cell reactivity to SARS-CoV-2 highlights the potential role of cellular immunity.^16,29,38^ Therefore, the Euroimmun anti-S1 measurements are not likely to provide a comprehensive measure of protective immunity following natural infection. In addition, our study population is not generalisable to all. Instead, it is representative of a workforce with likely high exposure, and low risk of severe COVID-19. Cohort 1 may have underestimated rates of infection because we were not able to recruit those who were in self-isolation at the peak of transmission, whilst cohort 2 may have overestimated rates of infection due to volunteer bias seeking testing.^39^ We do not expect either of these to confound our key findings. The sample size limited stratification of inter-individual heterogeneity, or identifying predictors of sero-reversion.

## Conclusions

Asymptomatic and pauci-symptomatic infection with SARS-CoV-2 elicits antibody responses to spike protein and to nuclear protein antigens in the vast majority, but with heterogeneity and differential temporal profiles. Anti-S1 antibodies measured by the Euroimmun assay have a shorter half-life, transition from high to lower levels of antibody production earlier and exhibit a greater reduction in antibody production rate, compared to anti-NP antibodies measured by the Roche assay. The important consequences of this are that, used alone, anti-S1 assays may underestimate past infection with implications for the application of this test for individual patient care and population level epidemiological surveys. Further work is required to evaluate the generalisability of these findings and investigate the determinants of heterogeneity in these antibody responses.

## Supporting information

Investigators

## Data Availability

The COVIDsortium Healthcare Workers consortium was prospectively designed to create a bioresource with high-dimensional sampling including viral PCR swabs, serology and PBMCs over an initial 20 weeks and pending 6-month and 1 year timepoints (study protocol has been published and is available online https://covid-consortium.com).
Applications for access to the individual participant de-identified data (including data dictionaries) and samples can be made to the access committee via an online application https://covid-consortium.com/application-for-samples/. Each application will be reviewed, with decisions to approve or reject an application for access made on the basis of (i) accordance with participant consent and alignment to the study objectives (ii) evidence for the capability of the applicant to undertake the specified research and (iii) availability of the requested samples.
The use of all samples and data will be limited to the approved application for access and stipulated in the material and data transfer agreements between participating sites and investigators requesting access.

## Footnotes

### Data Sharing Statement

The COVIDsortium Healthcare Workers consortium was prospectively designed to create a bioresource with high-dimensional sampling including viral PCR swabs, serology and PBMCs over an initial 20 weeks and pending 6-month and 1 year timepoints (study protocol has been published and is available online https://covid-consortium.com).^23^ Applications for access to the individual participant de-identified data (including data dictionaries) and samples can be made to the access committee via an online application https://covid-consortium.com/application-for-samples/. Each application will be reviewed, with decisions to approve or reject an application for access made on the basis of (i) accordance with participant consent and alignment to the study objectives (ii) evidence for the capability of the applicant to undertake the specified research and (iii) availability of the requested samples. The use of all samples and data will be limited to the approved application for access and stipulated in the material and data transfer agreements between participating sites and investigators requesting access.

