## Supplementary Figures for "Characterising heterogeneity and sero-reversion in antibody responses to mild SARS⍰CoV-2 infection: a cohort study using time series analysis and mechanistic modelling"

#### Supplementary Table 1. Baseline characteristics of anti-S1 seroconverters

| **Characteristic** | **Overall, N = 142** | **Anti-S1 Seroreversion, N = 32^1^** | **No anti-S1 seroreversion, N = 110^1^** |
| --- | --- | --- | --- |
| Age | 38 (30, 49) | 35 (32, 46) | 38 (30, 50) |
| Male (vs. Female) | 49 (35%) | 9 (28%) | 40 (36%) |
| BAME (vs. White) | 42 (31%) | 6 (20%) | 36 (34%) |
| Unknown | 7 | 2 | 5 |
| Case-defining symptoms (Yes vs. No) | 63 (44%) | 10 (31%) | 53 (48%) |
| Peak anti-S1 | 5.19 (2.85, 7.38) | 2.30 (1.66, 3.30) | 5.79 (3.77, 8.25) |
| Peak anti-NP | 63 (19, 107) | 18 (9, 37) | 79 (34, 115) |
| Unknown | 7 | 5 | 2 |
| ^1^Statistics presented: median (IQR); n (%) | | | |

### Supplementary Table 2. Peak anti-S1 response

|  | **Univariable** | | | | **Multivariable** | | |
| --- | --- | --- | --- | --- | --- | --- | --- |
| **Characteristic** | **N** | **Beta** | **95% CI^1^** | **p-value** | **Beta** | **95% CI^1^** | **p-value** |
| Age (per year increase) | 142 | 0.06 | 0.01, 0.10 | 0.009 | 0.05 | 0.01, 0.09 | 0.021 |
| Male (vs. Female) | 142 | 0.50 | -0.51, 1.5 | 0.3 | 0.09 | -0.93, 1.1 | 0.9 |
| BAME (vs. White) | 135 | 1.1 | 0.06, 2.2 | 0.039 | 1.00 | -0.04, 2.0 | 0.058 |
| Case-defining symptoms (Yes vs. No) | 142 | 0.78 | -0.19, 1.7 | 0.11 | 0.82 | -0.14, 1.8 | 0.094 |
| ^1^CI = Confidence Interval | | | | | | | |

### Supplementary Table 3. Peak anti-NP response

|  | **Univariable** | | | | **Multivariable** | | |
| --- | --- | --- | --- | --- | --- | --- | --- |
| **Characteristic** | **N** | **Beta** | **95% CI^1^** | **p-value** | **Beta** | **95% CI^1^** | **p-value** |
| Age (per year increase) | 150 | 0.61 | -0.09, 1.3 | 0.085 | 0.50 | -0.22, 1.2 | 0.2 |
| Male (vs. Female) | 149 | 2.4 | -14, 19 | 0.8 | 0.05 | -17, 17 | >0.9 |
| BAME (vs. White) | 144 | 23 | 6.0, 40 | 0.008 | 22 | 4.9, 39 | 0.012 |
| Case-defining symptoms (Yes vs. No) | 150 | 1.5 | -14, 17 | 0.9 | -0.79 | -16, 15 | >0.9 |
| ^1^CI = Confidence Interval | | | | | | | |

### Supplementary Table 4. Anti-S1 time to sero‑reversion

|  | **Univariable** | | | | **Multivariable** | | |
| --- | --- | --- | --- | --- | --- | --- | --- |
| **Characteristic** | **N** | **HR^1^** | **95% CI^1^** | **p-value** | **HR^1^** | **95% CI^1^** | **p-value** |
| Age (per year increase) | 142 | 0.98 | 0.95, 1.01 | 0.2 | 1.01 | 0.97, 1.05 | 0.6 |
| Male (vs. Female) | 142 | 0.93 | 0.43, 2.01 | 0.9 | 1.06 | 0.46, 2.45 | 0.9 |
| BAME (vs. White) | 135 | 0.58 | 0.24, 1.43 | 0.2 | 1.03 | 0.41, 2.64 | >0.9 |
| Case-defining symptoms (Yes vs. No) | 142 | 0.65 | 0.30, 1.38 | 0.3 | 0.66 | 0.30, 1.46 | 0.3 |
| Peak anti-S1 (per unit increase) | 142 | 0.41 | 0.30, 0.57 | <0.001 | 0.42 | 0.30, 0.58 | <0.001 |
| ^1^HR = Hazard Ratio, CI = Confidence Interval | | | | | | | |

#### Supplementary Figure 1. Relationship between antibody levels as measured by Euroimmun and Roche assays, and antibody dilution series.

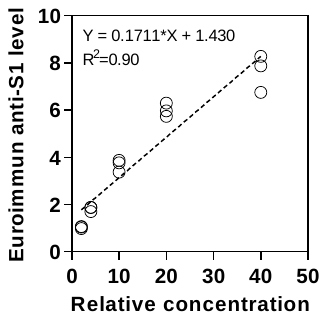

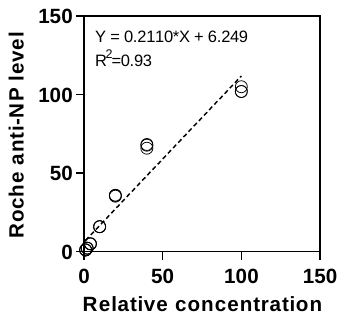

The measurement of antibody levels for each assay is shown across a range of serial dilutions.

#### Supplementary Figure 2. Mathematical model simulations of effect of selected parameters in changes to antibody levels.

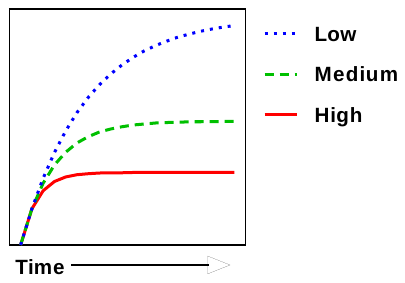

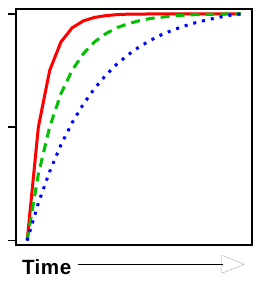

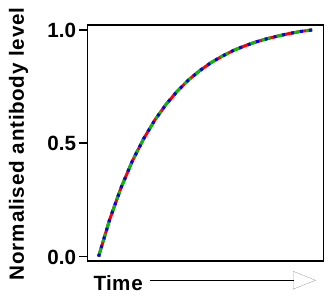

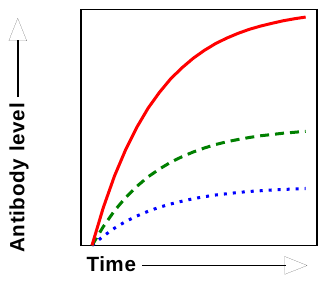

**A**

**B**

**C**

**D**

Variable **clearance** rate

Constant production rate

Variable **production** rate

Constant clearance rate

Increasing rates of antibody production at a constant rate of clearance leads higher antibody levels (A) but identical temporal profiles of antibody levels normalised to the peak (C). Increasing rates of clearance at a constant rate of production lead to lower antibody levels (B), but shorter time to peak antibody levels (D).

#### Supplementary Figure 3. Individual participant level serology data.

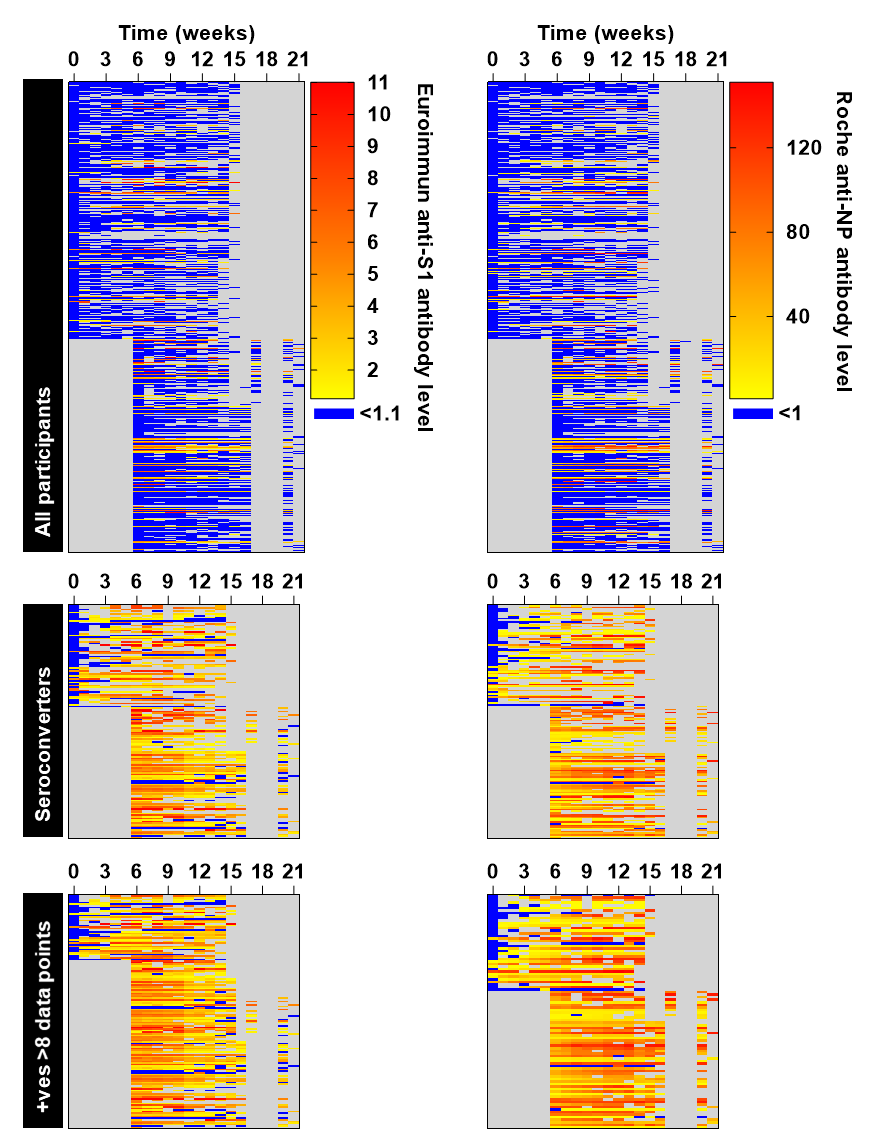

Heatmaps show antibody levels in each assay and missingness (grey) for individual participants over 0-21 weeks, for all participants (top row), all participants who were seropositive by at least one assay at any time point (middle row), and all such participants who had >8 data points available (bottom row), used for the mathematical modelling.

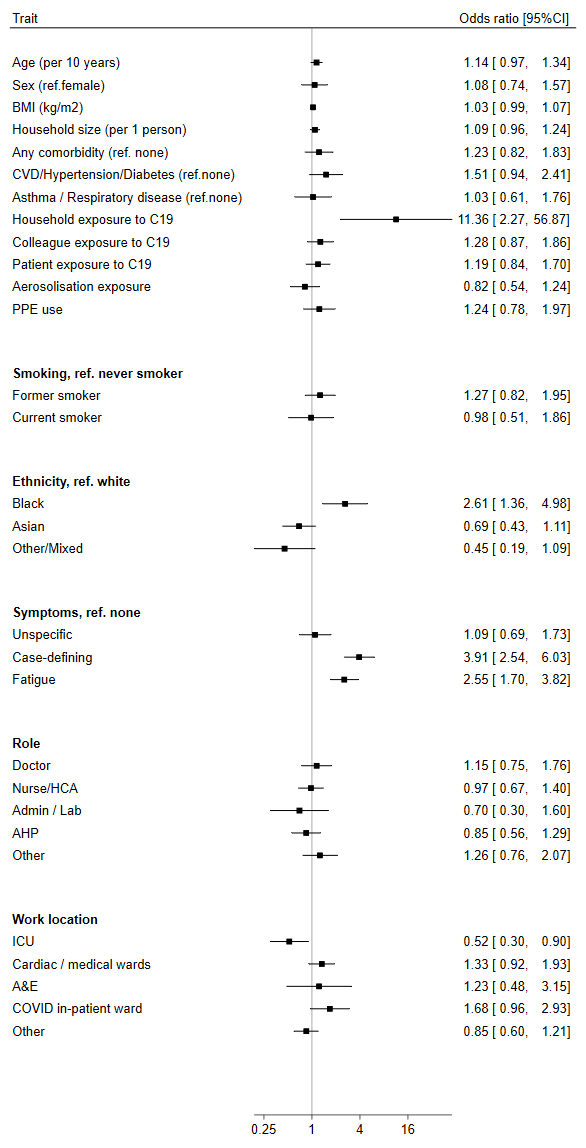

#### Supplementary Figure 4. Association of seropositivity to SARS-CoV-2 with demographic, clinical and exposure factors.

Odds ratios of selected demographic variables for seropositivity defined a participants who were seropositive at any time point using either assay across the study period

#### Supplementary Figure 5. Correlation of peak antibody responses in Euroimmun anti-S1 and Roche anti-NP assays

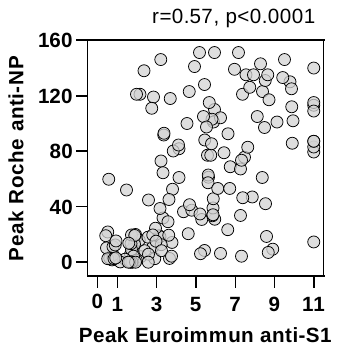

Peak antibody levels in each assay are shown for all participants who were seropositive by either assay at any time point, with Spearman rank correlation.

#### Supplementary Figure 6. Correlation in antibody levels stratified by time from infection.

**A**

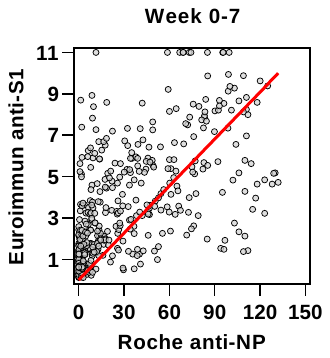

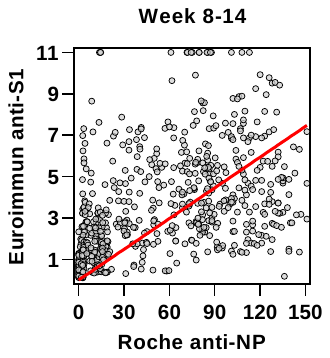

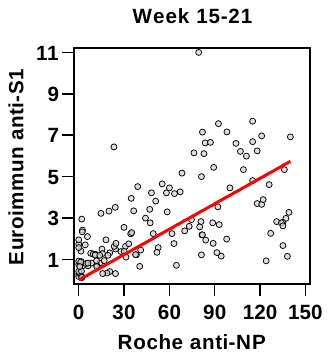

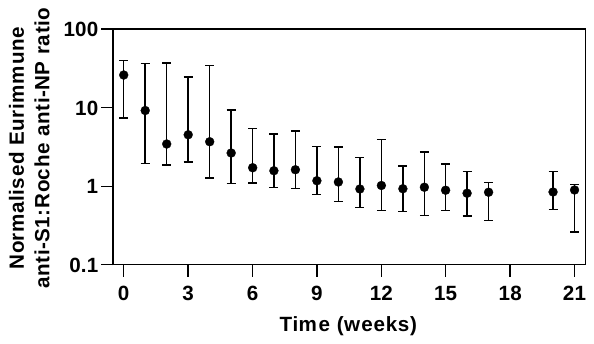

**B**

**(A)** For all participants who were seropositive by either assay at any time point, antibody levels from each assay are compared in three time intervals weeks 0-7 (left), weeks 8‑14 (centre) and weeks 15-21 (right). Red line shows the linear regression. **(B)** The normalised (by maximum value for each assay) ratio of Euroimmun antiS1 to Roche anti-NP by study week, showing median and interquartile range.

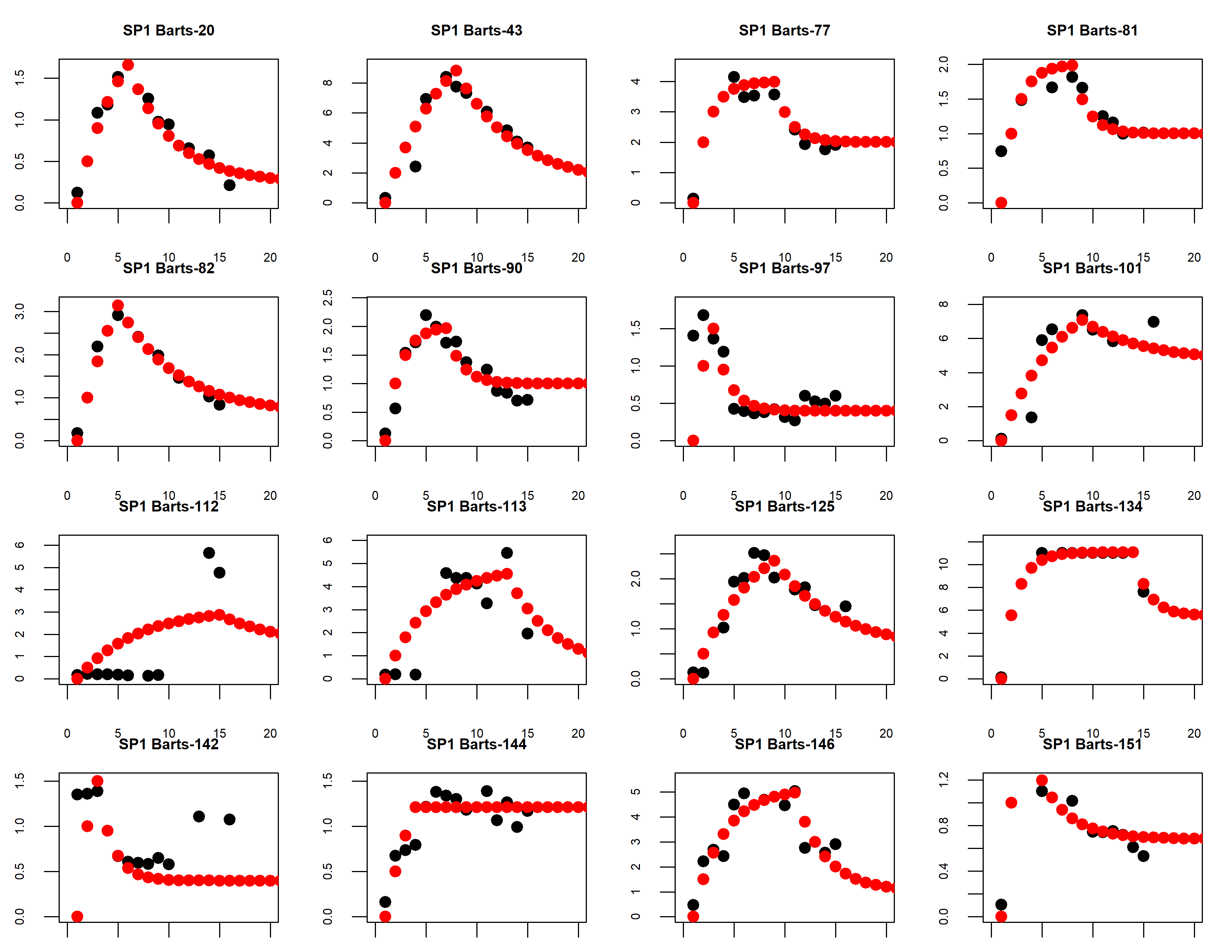

#### Supplementary Figure 7. Model fit to individual participant data

**A**

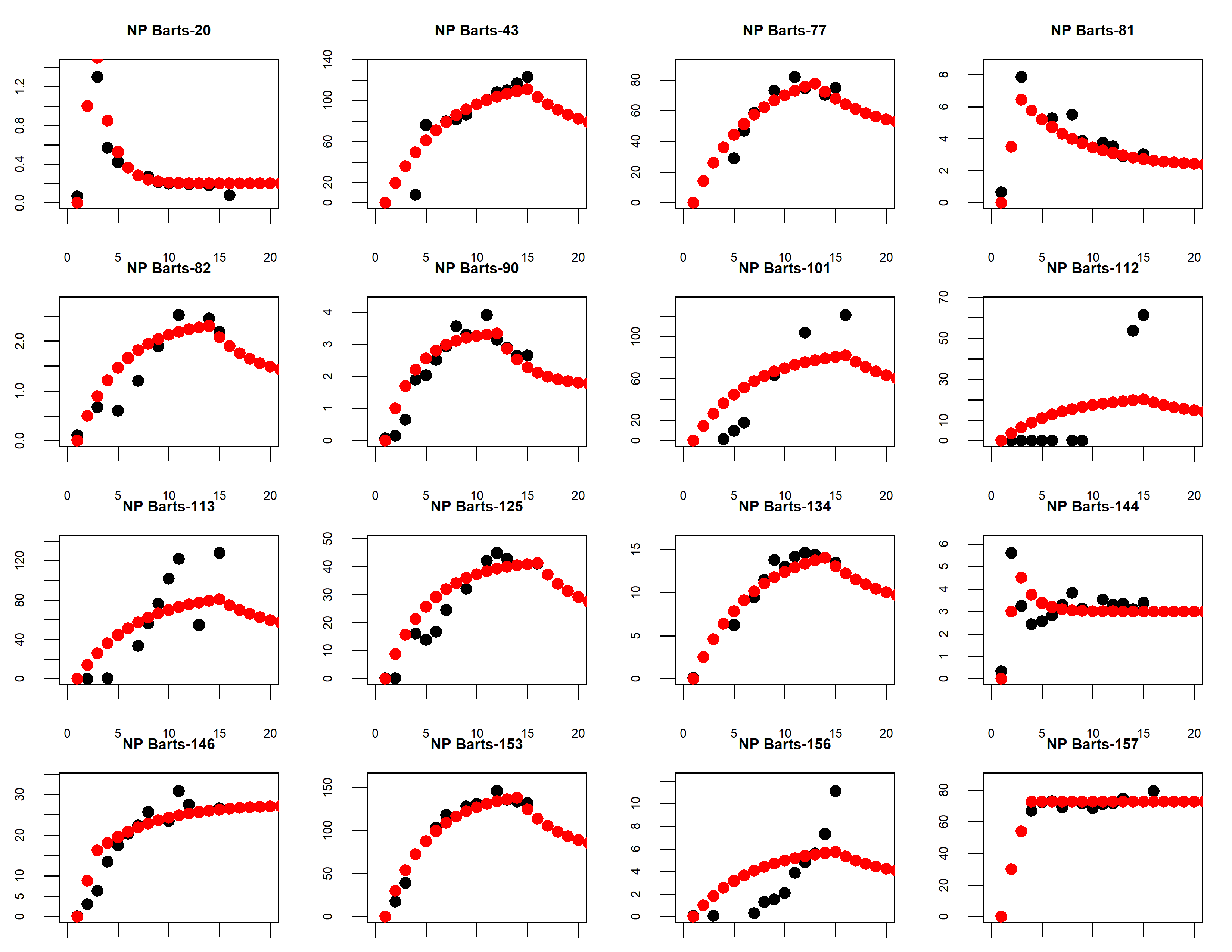

**B**

Representative models (red) fit to serology data (black) for **(A)** Euroimmun anti-S1 measurments, and **(B)** Roche anti‑NP measurements in 16 randomly selected individuals from participants who were seropositive with either assay at any time point, and had >8 serology data points available.

#### Supplementary Figure 8. Mathematical modelling of kinetics of circulating anti-S1 and anti-NP antibodies in baseline seronegative participants.

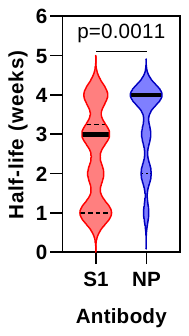

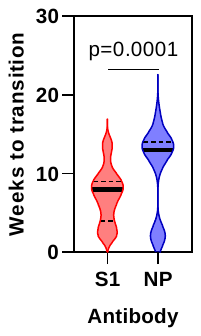

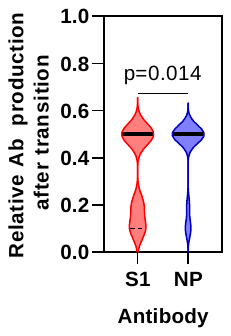

**C**

**B**

**A**

Best fit model parameters for **(A)** half-life of antibody clearance, **(B)** time to transition point of lower antibody production **(C)** and relative reduction in antibody production following this transition point, for individual baseline seronegative participants who subsequently seroconvert (N=50 for anti‑S1 serology, and 44 for anti‑NP serology; p values derived from 2‑tail Mann Whitney tests).
