## Supplementary material for "Characterising heterogeneity and sero-reversion in antibody responses to mild SARS⍰CoV-2 infection: a cohort study using time series analysis and mechanistic modelling": Investigators

**“COVIDsortium Investigators”**

Hakam Abbass, Aderonke Abiodun, Mashael Alfarih, Zoe Alldis, Daniel M Altmann, Mervyn Andiapen, Jessica Artico, Joao Augusto, Georgina L Baca, Anish Bhuva, Alex Boulter, Ruth Bowles , Rosemary J Boyton, Olivia Bracken, Timothy Brooks, Natalie Bullock, Gabriella Captur, Benny Chain, Nicola Champion, Carmen Chan, Jorge Couto de Sousa, Xose Couto-Parada , Marie-Teresa Cutino-Moguel, Rhodri H Davies, Keenan Dieobi-Anene, Karen Feehan, Malcolm Finlay, Marianna Fontana, Nasim Forooghi, Joseph M Gibbons, Derek Gilroy, Peter Griffiths, Rishi K Gupta, Matt Hamblin, Lauren M Hickling, Aroon D Hingorani, Lee Howes, Ivie Itua, Victor Jardim, Melanie Jensen, Meleri Jones, George Joy, Vikas Kapil, Jonathan Lambourne, WY Jason Lee, Mala K Maini, Vineela Mandadapu, Charlotte Manisty, Aine McKnight, Katia Menacho Medina, Celina Mfuko, Oliver Mitchelmore, James C Moon, Mahdad Noursadeghi, Ben O’Brien , Ben Ollivere, Corinna Pade, Susana Palma, Kush Patel, Ruth Parker, Brian Piniera, Alicja Rapala, Amy Richards, Mathew Robathan, Genine Sambile, Amanda Semper, Andreas Seraphim, Angelique Smit, Michelle Sugimoto, George D Thornton, Thomas A. Treibel, Arthur Tucker, Ana Valdes, Jessry Veerapen, Mohit Vijayakumar, Timothy Warner, Sophie Welch, Dylan Williams, Theresa Wodehouse , Lucinda Wynne, and Dan Zahedi.

**Public Health England National Infection Service Investigators**

Joanna Bacon, Daniel Bailey, Debbie Blick, Abbie Bown, Tim Brooks, Matthew Catton, Melanie Clifford, Mollie Curran-French, Silvia D’Arcangelo, Owen Daykin-Pont, Charlotte Dixon, Phoebe Do Carmo Silva, Ellie Drinkwater, Ross Fothergill, Harriet Garlant, Rachel Halkerston, Robin Hanson, James Hardy, Alexander Hargreaves, Jacqueline Hewson, Charlotte Hind, Emma Hobbs, Leah Johal, Jessica Jones, Deborah Lister, Adam Mabbutt, Bethany Martin, Lara Mason, Joanna McGlashan, Isobel Miles, Gloria Mongelli, Christopher Moon, Taalia Morgan, Alexander Morrison, Anna Moy, Joshua Nelthorpe-Cowne, Ashley Otter, Ros Packer, Jordan Pascoe, Prem Perumal, Adam Roberts, April Roberts, Cathy Rowe, Amanda Semper, Lauren Setterfield, Sara Speight, Deen Qureshi, Stephen Taylor, Stephen Thomas, Sian Tiley, Rosie Watts, Clare Wilson, and Charlotte Woolley.

**Public Health England data entry team**

Frances Alexander, Sarah Belcher, Collette Biggs, Cathy Collins, Ant Crook, Lisa Crook, Helen Davies, Melanie Davison, Amy Hankins, Jennifer Logue, Debbie Mason, Helen Philpott, Julia Sung, Cat Wood
